# Early estimates of the incidence trend and the reproductive number of the monkeypox epidemic in Brazil

**DOI:** 10.1101/2022.08.16.22278806

**Authors:** Isaac N. Schrarstzhaupt, Mellanie Fontes-Dutra, Fredi Alexander Diaz-Quijano

## Abstract

**Objectives:** We aimed to calculate the weekly growth of the incidence and the effective reproductive number (Rt) of the 2022 Monkeypox epidemic during its introduction in Brazil.

**Methods:** We described the case distribution in the country and calculated the incidence trend and the Rt in the four geographical states with the highest case reports. By using two regression approaches, count model and the Prais-Winsten, we calculated the relative incidence increase. Moreover, we estimated the Rt for the period between the 24th and the 50th days after the first official report, using a serial interval reported in another population and two alternative values (**±** 3 days).

**Results:** Up to August 22, 3.896 Monkeypox cases were confirmed in Brazil. The weekly incidence increases were between 37.5% (95% CI: 20.7% -56,6%) and 82.1% (95% CI: 59.5% -107.8%), and all estimates of Rt were statistically higher than 1 in the four states analyzed.

**Conclusion:** The Monkeypox outbreak in Brazil is a significant public health emergency that requires coordinated public health strategies such as testing, contact tracing, and vaccination.

## INTRODUCTION

In July of 2022, the World Health Organization declared that the monkeypox outbreak was a public health emergency of international concern (1). Up until August 22, 2022 there were 43.583 confirmed cases and 9 deaths worldwide (2). In Brazil, the first case was confirmed on June 8, 2022, increasing to 3.896 confirmed cases until August 22, 2022 (2). Brazil also registered one Monkeypox death, on July 29, 2022, in the state of Minas Gerais (MG).

Although this outbreak is unprecedented, Monkeypox is not a new disease. The virus was discovered in 1959 and it is an endemic disease in Central and West Africa (3,4). In addition, outbreaks were reported in non-endemic countries, such as the United States of America in 2003 and the United Kingdom in 2018 (5,6). Outbreaks were also reported in the Democratic Republic of the Congo, where Monkeypox is endemic, in 2013(7). Data on literature prior to the aforementioned outbreaks have warned about the risk of reintroduction of Monkeypox virus in the human population, once the virus might infect susceptible animals (8). Moreover, the additional protection by smallpox vaccine decreases over time in the population and, as the vaccine campaigns against smallpox were interrupted with the eradication of this disease, younger people were not immunized against this disease (9).

Incidence trends and reproductive number estimates are important to recognize the potential extension of an outbreak and to justify public health interventions (3). In this work, we aimed to estimate the weekly incidence growth and the effective reproductive number (Rt) of the 2022 Monkeypox epidemic during its introduction in Brazil.

## METHODS

This was an ecological study based on surveillance data of the epidemiological reports posted by the Brazilian National Health Ministry (10). With this data, we created a database with all of the new and accumulated cases and deaths (11). We focused on the four states with more than 150 cases up to August 22, 2022. These states are SP (São Paulo), RJ (Rio de Janeiro), MG and Goiás, which geographically includes the Federal District of Brasília (GO-FD).

The Brazilian Health Ministry defines a suspected Monkeypox case as a case that shows one or more of the following criteria(12):

- Prolonged exposure with no respiratory protection OR direct physical contact, including sexual contact, with multiple and/or unknown partner in the 21 days before the marks and/or symptoms’ onset;
- Prolonged exposure with no respiratory protection OR close contact history, including sexual, with suspected or confirmed Monkeypox case in the 21 days before the marks and/or symptoms’ onset;
- Contact with contaminated materials like sheets and bathroom towels or common use utensils that belong to a suspected or confirmed Monkeypox case in the 21 days before the marks and/or symptoms’ onset;
- Health workers without proper use of protection equipment with history of contact with suspected or confirmed Monkeypox case in the 21 days before the marks and/or symptoms’ onset.

Confirmed cases are defined by a suspected case with positive PCR result for the Monkeypox virus (MPXV)(12).

Considering the confirmed cases, we performed two analyses: first we estimated the weekly growth of the incidence of Monkeypox, starting one day after the first case reporting in the corresponding state. After grouping the cases by week, we used two regression approaches: first, we used count models of Poisson and negative binomial regression and reported the last one when the alpha term differed significantly from zero (13,14). We also applied the Prais-Winsten regression to the natural logarithm of the weekly case count (15,16). We reported the relative incidence increase calculated as 1-*e*^*β*^, where *β* is the regression coefficient. We chose to use both regressions to ensure that we have a consistent result on the weekly growth estimates.

Additionally, we estimated the Rt by using the “estimate_R” function contained in the “EpiEstim” package of the R programming language in each of these four states (17,18,19). We used the “parametric_si” configuration of the package (script available as supplementary material). One of the parameters that we need to estimate the Rt is the serial interval (SI) of the disease. The SI consists of looking at a pair of cases and extracting the amount of days between the day that the disease installs itself on the primary case and the day that it installs itself on the secondary case. Since the microdata of Monkeypox cases in Brazil are unavailable, we do not have the local SI data, so we used a value recently calculated in the United Kingdom (20). This SI was obtained from 17 pairs of cases (primary -secondary) of the United Kingdom. The mean SI calculated in this study is 9.8 days, and the standard deviation is 9.1 days (20). Since SI can be different from one location to another, we also calculated the Rt for two more SI values defined by subtracting and adding three days to the reference (6.8 and 12.8 days, respectively) (20).

The Rt is estimated on weekly sliding windows, given by the parameters “t_start” and “t_end”. The cases first started to be officially notified in Brazil on June 26, 2022, and we calculated the daily Rt oscillation from the 24th day after that day. We also calculated the mean Rt with the same method, using the “estimate_R” function, but with the parameter “t_start” as 24 and the parameter “t_end” as 50, as a summary estimate for each state.

This study used public data, and the analysis was performed using R (version 4.2.1) and Stata (version 17.0, Stata Corp LP, College Station, TX, USA)

## RESULTS

The 3.896 confirmed Monkeypox cases in Brazil up to August 22, 2022, almost doubled when compared to the 2.004 cases that were notified up to August 5, 2022. The state with the most reported cases was SP, followed by RJ and GO-FD (Figures 1 and 2). However, the highest relative increase of incidence was observed in GO-FD, being 74.7% per week using the count model and 82.1% using the Prais-Winsten regression. In all states, the increasing trend was statistically significant with both regression approaches (Table).

**Table 1.** Relative incidence increase and reproductive number of monkeypox epidemic in the most affected states of Brazil.

**Table. Relative incidence increase and reproductive number of monkeypox epidemic in the most affected states of Brazil.**
| State | Weekly incidence increase (95% CI) |  | Rt - mean (95% CI) according serial interval (SI) |  |  |
| --- | --- | --- | --- | --- | --- |
|  | Count model* | Prais-Winsten | SI: 9.8 days | Sensitivity analysis: |  |
|  |  |  |  | SI: 6.8 days | SI: 12.8 days |
| GO-FD† | 74.7% (52.5% - 100.2%) | 82.1% (59.5% - 107.8%) | 2.07 (1.98 – 2.16) | 1.73 (1.66 – 1.80) | 2.42 (2.31 – 2.52) |
| SP‡ | 45.5% (21.9% - 73.7%) | 45.6% (15.5% - 83.6%) | 1.70 (1.68 – 1.72) | 1.46 (1.45 – 1.48) | 1.94 (1.92 – 1.96) |
| RJ§ | 37.5% (20.7% - 56.6%) | 40% (18.4% - 65.6%) | 1.65 (1.61 – 1.70) | 1.44 (1.40 – 1.48) | 1.87 (1.82 – 1.92) |
| MG¶ | 41.5% (31.7% - 52%) | 42.9% (39% - 46.9%) | 1.64 (1.57 – 1.72) | 1.43 (1.37 – 1.50) | 1.86 (1.78 – 1.94) |

\*Estimate for MG was obtained using Poisson, and for the other states using negative binomial regress (p-value <0.001 of alpha=0).
† GO-FD: the state of Goiás plus Federal District (Brasília);; ‡SP: São Paulo; §RJ: Rio de Janeiro; ¶MG: Minas Geraí

**Figure 1:**
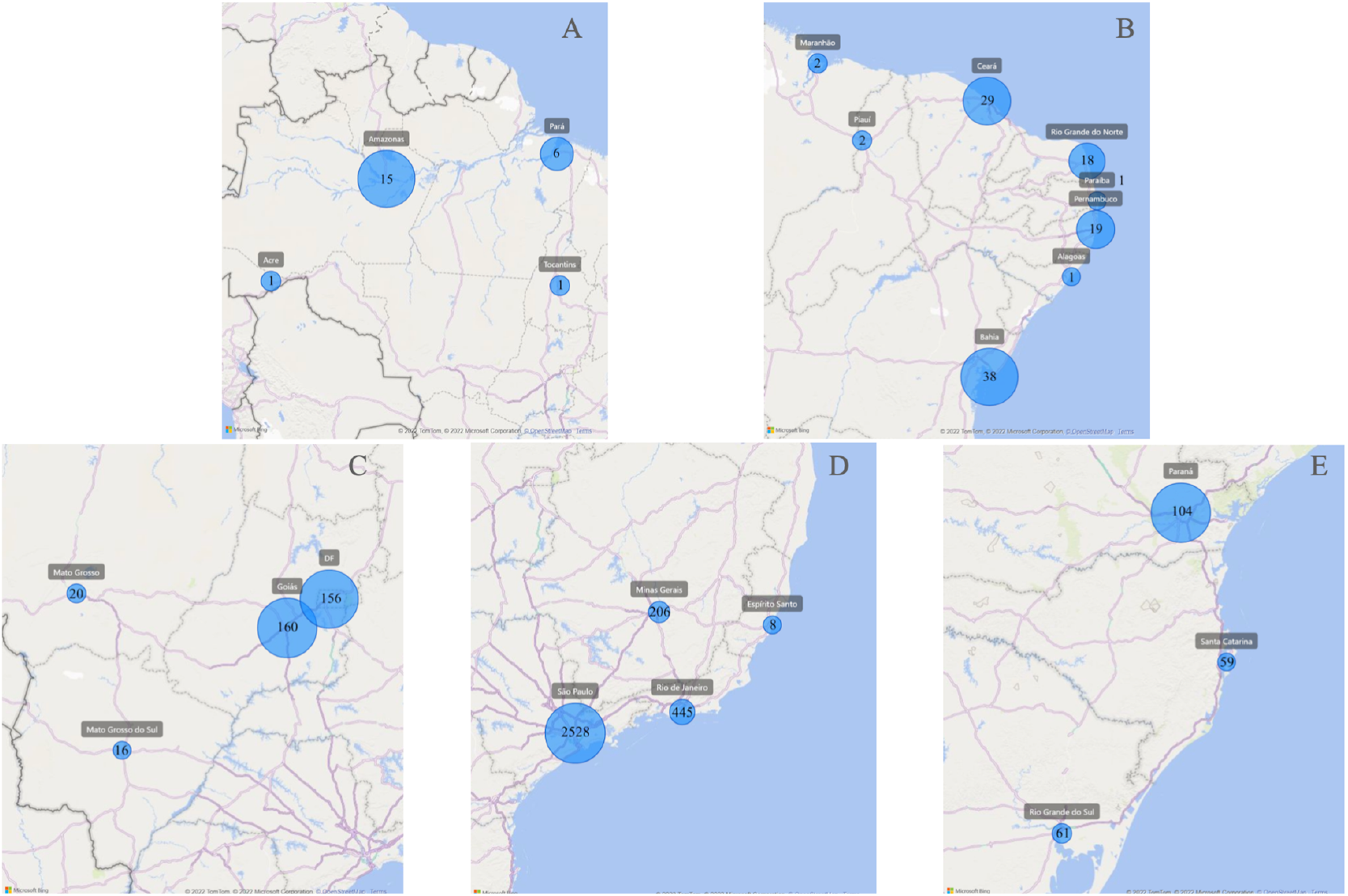
Brazil maps with Monkeypox incidence by region: North (A), Northeast (B), Center West (C), Southeast (D) and South (E)

**Figure 2:**
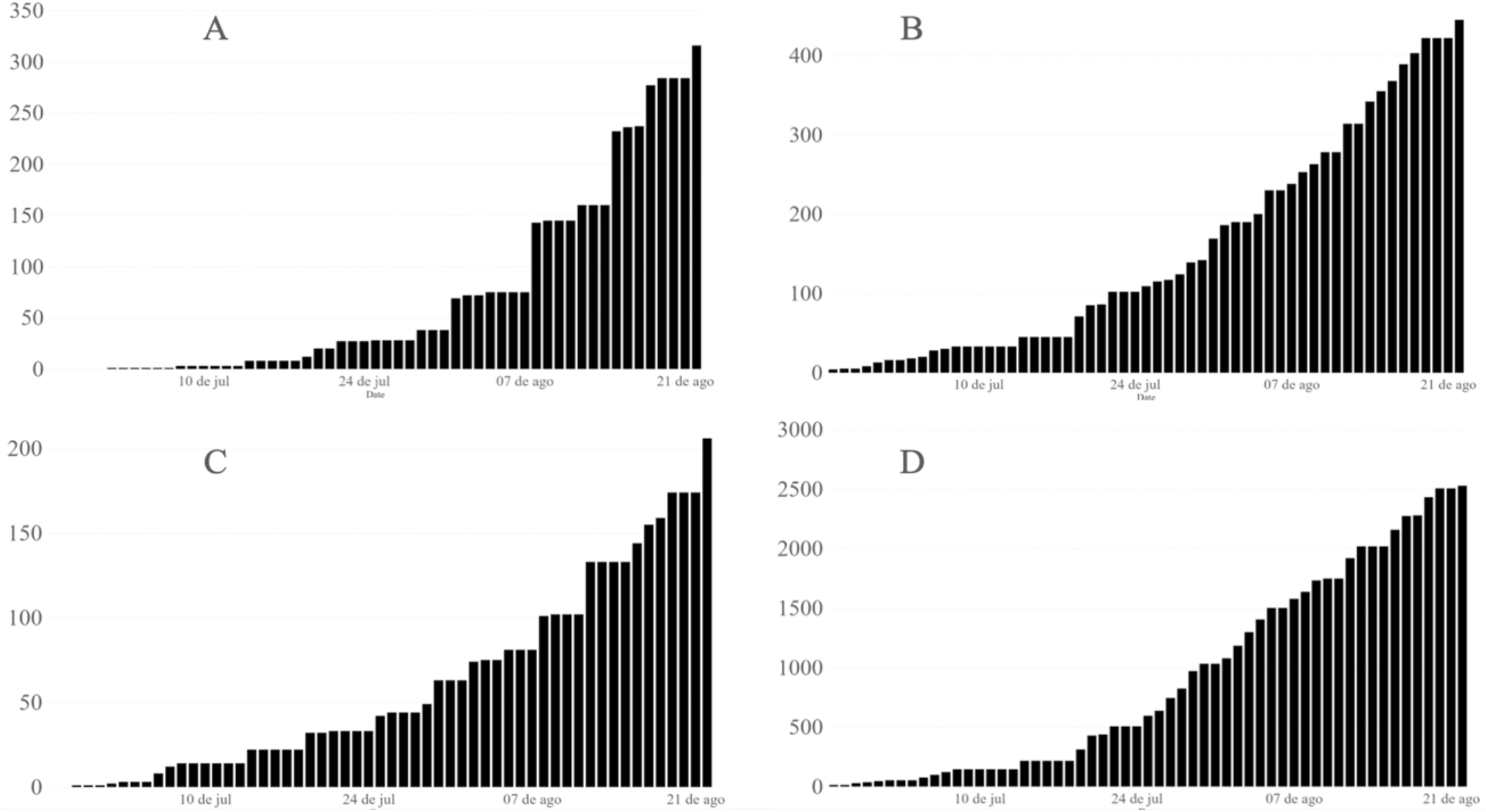
Epidemic curve of the four analyzed states: GO-FD (A), RJ (B), MG (C) and SP (D)

The Rt was also higher in GO-FD, with 2.07 (95% CI: 1.98 -2.16), followed by SP with 1.70 (1.68 -1.72), RJ with 1.65 (1.61,1.70) and MG with 1.64 (1.57,1.72). The Rt, estimated after the 24th day of the outbreak with a SI of 9.8 days, oscillates between 1.86 and 2.62 in GO-FD, 1.54 and 2.00 in RJ, 1.56 and 1.77 in MG and 1.48 and 2.16 in SP (Figure 3). With SIs of 6.8 days and 12.8 days (−3 and +3 days) all the Rt estimates stayed significantly above 1 (Figure 4).

**Figure 3:**
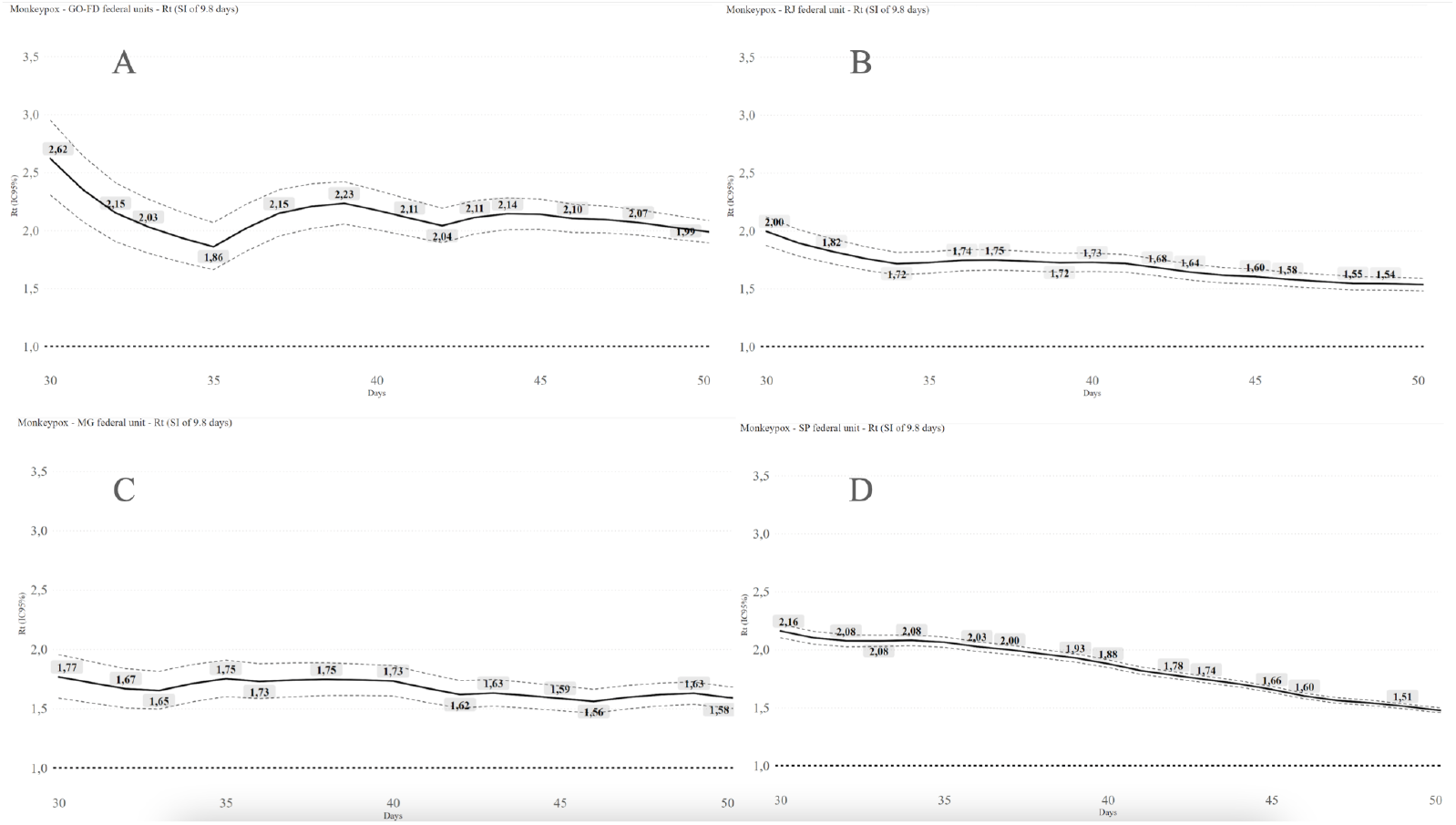
Rt estimates from GO-FD (A), RJ (B), MG (C) and SP (D) with an SI of 9.8 days

**Figure 4:**
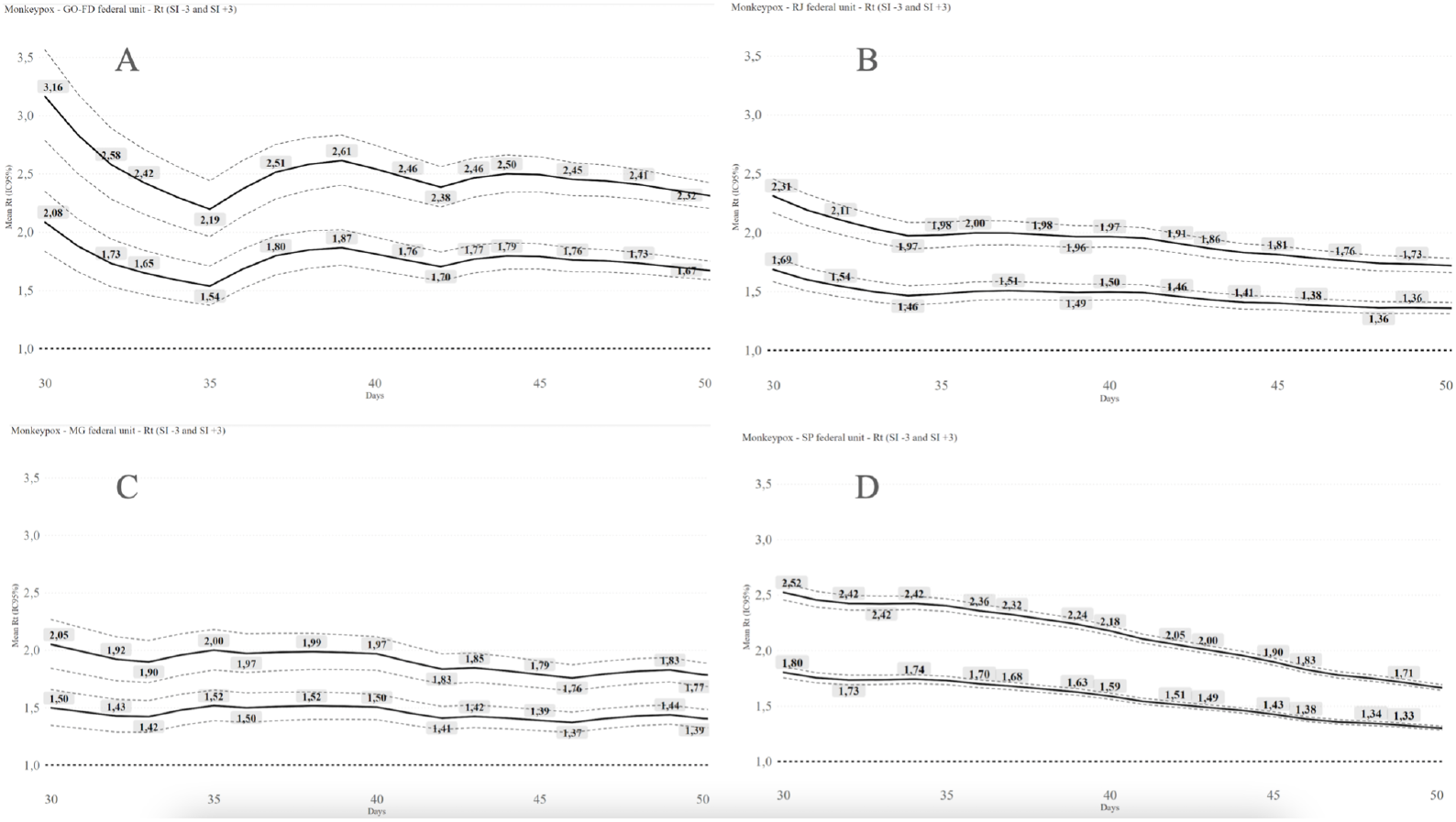
Rt estimates from GO-FD (A), RJ (B), MG (C) and SP (D) with SIs of 6.8 and 12.8 days (−3 and +3)

## DISCUSSION

In the present study, we illustrated a significant increase in confirmed Monkeypox cases in Brazil up to August 22, 2022. We are also observing a similar pattern in other states (data not shown), like Paraná and Rio Grande do Sul, even with lower numbers of cases (104 and 61, respectively). The values of Rt estimated for four states are considerably high and may be comparable with some calculated for COVID-19 (13). However, it is important to highlight that these two infectious agents have significantly different profiles of transmission. Monkeypox transmission appears to mainly occur by direct contact in the current outbreak, which can be related to a high viral load on skin lesions (22,23). Moreover, there is evidence of a lesion staying positive on the PCR test for up to 21 days (24). We hypothesize that a prolonged infectious period could explain high Rt values even if the ways of transmission were limited.

This Rt is also consistent with other findings. Kwok et al. (2022) estimated a Ro of 1.6 (1.5-1.7) in England, 1.4 (1.2-1.6) in Portugal and 1.8 (1.7-2.0) in Spain(20). Despite the differences among these populations, these data indicate the increasing transmission of Monkeypox in human populations during the current outbreak. Therefore, it is urgent to identify and interrupt the main transmission routes.

Our study had some methodological limitations. The sample size was limited and Rt can be unstable during the early stages of the epidemic (25). Therefore, we restricted our analysis to four states and the Rt was estimated after the 24th day of the first official case notification. Another limitation is the potential delay in both notification and testing, affecting the real-time monitoring of the incidence.

We estimated the Rt with a preset SI obtained from another population, from the United Kingdom, since the Brazilian microdata was unavailable (20). However, we believe this value could be compatible with that expected in Brazil since it was recently calculated in the context of the current epidemic. On the other hand, the sensitivity analysis, with alternative values of SI, supports robust conclusions regarding the lack of transmission control. In that sense, our Rt estimates correlated well with the observed upward trends across the states, which give a reference of the potential transmissibility of Monkeypox in Brazil.

Our results highlight the need for effective public health measures such as testing, contact tracing and isolation of the confirmed cases. There is also a need for smallpox vaccination which could protect against Monkeypox (26), in spite of the fact that we do not have a specific Monkeypox vaccine at this moment. It also reinforces the importance of surveillance and more detailed public data to ensure that more analysis can be made. The weekly growth rates might be related to long infectious periods of the Monkeypox disease. If this hypothesis is confirmed with further studies, it could guide assertive public health interventions, such as prolonged isolation, to counteract the current epidemic.

## Supporting information

R Script for the Rt estimative

## Data Availability

All data produced in the present study are available upon reasonable request to the authors

https://github.com/isaacdata/Rt_monkeypox

## Acknowledgements

We would like to acknowledge Leonardo Medeiros for collecting the data on the official daily postings of the Brazilian Health Ministry and Leonardo Bastos for helping with R packages and references.

## REFERENCES

1. WHO Director-General’s statement at the press conference following IHR Emergency Committee regarding the multi-country outbreak of monkeypox - 23 July 2022 [Internet]. [citado 12 de agosto de 2022]. Disponível em: https://www.who.int/director-general/speeches/detail/who-director-general-s-statement-on-the-press-conference-following-IHR-emergency-committee-regarding-the-multi--country-outbreak-of-monkeypox--23-july-2022

2. Global.health | Map [Internet]. [citado 12 de agosto de 2022]. Disponível em: https://map.monkeypox.global.health/country

3. Magnus P von, Andersen EK, Petersen KB, Birch-Andersen A. A Pox-Like Disease in Cynomolgus Monkeys. Acta Pathol Microbiol Scand. 1959;46(2):156–76.

4. Hoff NA, Doshi RH, Colwell B, Kebela-Illunga B, Mukadi P, Mossoko M, et al. Evolution of a Disease Surveillance System: An Increase in Reporting of Human Monkeypox Disease in the Democratic Republic of the Congo, 2001–2013. Int J Trop Dis Health. 2017;25(2):IJTDH.35885.

5. Centers for Disease Control and Prevention (CDC). Update: multistate outbreak of monkeypox--Illinois, Indiana, Kansas, Missouri, Ohio, and Wisconsin, 2003. MMWR Morb Mortal Wkly Rep. 11 de julho de 2003;52(27):642–6.

6. Vaughan A, Aarons E, Astbury J, Balasegaram S, Beadsworth M, Beck CR, et al. Two cases of monkeypox imported to the United Kingdom, September 2018. Euro Surveill Bull Eur Sur Mal Transm Eur Commun Dis Bull. setembro de 2018;23(38).

7. Nolen LD, Osadebe L, Katomba J, Likofata J, Mukadi D, Monroe B, et al. Extended Human-to-Human Transmission during a Monkeypox Outbreak in the Democratic Republic of the Congo. Emerg Infect Dis. junho de 2016;22(6):1014–21.

8. Fine PEM, Jezek Z, Grab B, Dixon H. The Transmission Potential of Monkeypox Virus in Human Populations. Int J Epidemiol. 1o de setembro de 1988;17(3):643–50.

9. Nguyen PY, Ajisegiri WS, Costantino V, Chughtai AA, MacIntyre CR. Reemergence of Human Monkeypox and Declining Population Immunity in the Context of Urbanization, Nigeria, 2017-2020. Emerg Infect Dis. abril de 2021;27(4).

10. Atualização dos Casos [Internet]. Ministério da Saúde. [citado 12 de agosto de 2022]. Disponível em: https://www.gov.br/saude/pt-br/composicao/svs/resposta-a-emergencias/coes/monkeypox/atualizacao-dos-casos

11. Schrarstzhaupt I. Rt_monkeypox [Internet]. 2022 [citado 18 de agosto de 2022]. Disponível em: https://github.com/isaacdata/Rt_monkeypox

12. Definição de Caso [Internet]. Ministério da Saúde. [citado 12 de agosto de 2022]. Disponível em: https://www.gov.br/saude/pt-br/assuntos/variola-dos-macacos/definicao-de-caso/definicao-de-caso

13. Hoffmann JP. Regression models for categorical, count, and related variables: an applied approach. Oakland: University of California Press; 2016.

14. Coxe S, West SG, Aiken LS. The Analysis of Count Data: A Gentle Introduction to Poisson Regression and Its Alternatives. J Pers Assess. 17 de fevereiro de 2009;91(2):121–36.

15. Antunes JLF, Cardoso MRA. Uso da análise de séries temporais em estudos epidemiológicos. Epidemiol E Serviços Saúde. setembro de 2015;24:565–76.

16. Prais SJ, Winsten CB. Trend estimators and serial correlation. Cowles Com Discuss Pap [Internet]. 24 de fevereiro de 1954;(Statistics No 383). Disponível em: https://cowles.yale.edu/sites/default/files/files/pub/cdp/s-0383.pdf

17. Cori [aut A, cre, Cauchemez S, Ferguson NM, Fraser C, Dahlqwist E, et al. EpiEstim: Estimate Time Varying Reproduction Numbers from Epidemic Curves [Internet]. 2021 [citado 18 de agosto de 2022]. Disponível em: https://CRAN.R-project.org/package=EpiEstim

18. R: The R Project for Statistical Computing [Internet]. [citado 18 de agosto de 2022]. Disponível em: https://www.r-project.org/

19. make_config function - RDocumentation [Internet]. [citado 12 de agosto de 2022]. Disponível em: https://www.rdocumentation.org/packages/EpiEstim/versions/2.2-4/topics/make_config

20. Kwok KO, Wei WI, Tang A, Shan Wong SY, Tang JW. Estimation of local transmissibility in the early phase of monkeypox epidemic in 2022. Clin Microbiol Infect [Internet]. 8 de julho de 2022 [citado 12 de agosto de 2022]; Disponível em: https://www.sciencedirect.com/science/article/pii/S1198743X22003408

21. Billah MA, Miah MM, Khan MN. Reproductive number of coronavirus: A systematic review and meta-analysis based on global level evidence. PloS One. 2020;15(11):e0242128.

22. Monkeypox [Internet]. [citado 14 de agosto de 2022]. Disponível em: https://www.who.int/news-room/fact-sheets/detail/monkeypox

23. Tarín-Vicente EJ, Alemany A, Agud-Dios M, Ubals M, Suñer C, Antón A, et al. Clinical presentation and virological assessment of confirmed human monkeypox virus cases in Spain: a prospective observational cohort study. The Lancet [Internet]. 8 de agosto de 2022 [citado 15 de agosto de 2022];0(0). Disponível em: https://www.thelancet.com/journals/lancet/article/PIIS0140-6736(22)01436-2/fulltext#seccestitle10

24. Thornhill JP, Barkati S, Walmsley S, Rockstroh J, Antinori A, Harrison LB, et al. Monkeypox Virus Infection in Humans across 16 Countries — April–June 2022. N Engl J Med. 21 de julho de 2022;0(0):null.

25. Cintrón-Arias A, Castillo-Chávez C, Bettencourt LMA, Lloyd AL, Banks HT. The estimation of the effective reproductive number from disease outbreak data. Math Biosci Eng. 1° de abril de 2009;6(2):261–82.

26. Edghill-Smith Y, Golding H, Manischewitz J, King LR, Scott D, Bray M, et al. Smallpox vaccine-induced antibodies are necessary and sufficient for protection against monkeypox virus. Nat Med. julho de 2005;11(7):740–7.

